# Study Protocol- The impact of social deprivation on development and progression of diabetic kidney disease

**DOI:** 10.1101/2024.04.24.24306283

**Authors:** Caoimhe Casey, Claire M Buckley, Patricia M Kearney, Matthew D Griffin, Sean F Dinneen, Tomás P Griffin

## Abstract

**Introduction:** Diabetes is one of the leading causes of chronic kidney disease. Social deprivation is recognised as a risk factor for complications of diabetes, including diabetic kidney disease. The effect of deprivation on rate of decline in renal function has not been explored in the Irish Health System to date. The objective of this study is to explore the association between social deprivation and the development/progression of diabetic kidney disease in a cohort of adults living with diabetes in Ireland.

**Methods and analysis:** This is a retrospective cohort study using an existing dataset of people living with diabetes who attended the diabetes centre at University Hospital Galway from 2012 to 2016. The variables included in this dataset include demographic variables, type and duration of diabetes, clinical variables such as medication use, blood pressure and BMI and laboratory data including creatinine, urine albumin to creatinine to ratio, haemoglobin A1c and lipids. This dataset will be updated with laboratory data until January 2023. Individual’s addresses will be used to calculate deprivation indices using the Pobal Haase Pratschke (HP) deprivation index. Rate of renal function decline will be calculated using linear mixed-effect models. The relationship between deprivation and renal function will be assessed using linear regression (absolute and relative rate of renal function decline based on eGFR) and logistic regression models (rapid vs. non-rapid decline).

**Ethics and dissemination:** Ethical approval has been granted by the clinical research ethics committee of Galway University Hospitals-Ref C.A. 2956. Results will be presented at conferences and published in peer review journals.

## Introduction

In 2021, 537 million adults aged 20-79 were estimated to live with diabetes worldwide, with projections suggesting an increase to 643 million by 2030 (1). Diabetes accounted for approximately 6.7 million deaths globally in 2021. Diabetes is one of the leading causes of chronic kidney disease (CKD) (2). The number of new cases of CKD due to type 2 diabetes increased worldwide from approximately 1.4 million in 1990 to 2.4 million in 2017 (2). There is an inverse association between CKD incidence and a country’s sociodemographic index (2). Mortality is increased in people with diabetic kidney disease (DKD) (3). Those with kidney disease and type 2 diabetes have a standardised mortality rate of 31.3%, compared to 11.5% in those with diabetes without kidney disease (3). CKD due to diabetes is defined as persistent albuminuria (an albumin-to-creatinine ratio >3mg/mmol), persistently reduced renal function (an estimated Glomerular Filtration Rate <60ml/min per 1.73m2) or both for greater than 3 months (4). Hypertension, dyslipidaemia, hyperglycaemia and smoking are some modifiable risk factors for DKD (5). Socioeconomic deprivation is associated with DKD through its influence on these risk factors, but also through other mechanisms such as access to healthcare and health literacy (6).

Socioeconomic deprivation, is characterized by restricted access to societal resources due to poverty, discrimination and other disadvantages (7), The prevalence of type 2 diabetes is linked to social deprivation, with a higher prevalence in deprived groups (8, 9), likely due to barriers to healthy living conditions, education, and behavioural factors imposed by economic constraints. The relationship between deprivation and type 1 diabetes is less clearly understood with studies reporting varied relationships ranging from no association (10), to an inverse association (11) to a positive association (12). Socioeconomic deprivation is also a risk factor for complications in both type 1 and type 2 diabetes including diabetic retinopathy (13, 14), cardiovascular disease (15, 16), diabetic foot ulceration and amputation (17) and mortality in type 1 and 2 diabetes (18–20). There is also significant evidence of an influence of social deprivation on development of DKD. The EURODIAB IDDM complications study showed a significant association between lower educational attainment and macroalbuminuria (21). Gonzales et al demonstrated in the UK that among people living with type 1 diabetes, those living in the most deprived circumstances had a hazard ratio of 2.92 for incident DKD compared to the least deprived. Similarly in type 2 diabetes, the most deprived had a hazard ratio of 1.39 for incident DKD compared to the least deprived (22).

Studies to date in Ireland on deprivation and diabetes have focussed on diabetes prevalence and predominantly use individual level markers of socioeconomic deprivation such as education and occupation (23–25). Educational attainment, a component used in determining deprivation status, has been shown to be inversely associated with the prevalence of multimorbidity in a cohort study in Ireland (23). Participants completed health and lifestyle questionnaires and attended for a physical exam. Multimorbidity was defined as the presence of two or more chronic diseases including diabetes. Educational attainment was ascertained from the questionnaire and divided into primary level or secondary level and above. O Connor et al looked at the determinants of undiagnosed and diagnosed diabetes and used social class (as defined by the European Socioeconomic Classification System), education and medical insurance as covariates (24). Insurance was classified into “private insurance” (paid for by the individual), means tested state assisted –“state insurance” and no insurance and results showed that those with private insurance were less likely to have a diagnosis of type 2 diabetes and those with who finished education at primary level were more likely to have a diagnosis of type 2 (24). Leahy et al used the Irish Longitudinal Study of Ageing (TILDA), a study on adults over 50 years in Ireland and looked at social class based on fathers’ occupation. Compared with professional/managerial occupations, belonging to the “manual” social class in childhood was associated with an increased risk of type 2 diabetes (25). Socioeconomic deprivation is associated with non-attendance at the Irish national diabetic retinopathy screening service (26). The most deprived quintile had approximately 12% higher non-attendance rates compared to the middle quintile. Deprivation is associated with an increased rate of admission to hospital for diabetes complications – diabetic ketoacidosis, renal complications, retinopathy, neuropathy, peripheral vascular disease and “other” complications (27). This was shown after adjusting for population density and medical-card (state insurance) coverage (27). Interestingly, a study of over 1000 people with type 1 and type 2 diabetes attending primary care showed no difference in haemoglobin A1c (HbA1c) values between deprivation categories, even after adjusting for whether the diabetes care was shared with secondary care or managed in primary care alone (28). However, this does not take into account that attendance rates may be affected by deprivation and those with higher HbA1c values may be less likely to attend for monitoring. Another study using the TILDA dataset, looked at risk factors for macro and micro vascular complications in type 2 diabetes (29) and showed that higher educational attainment was associated with a lower likelihood of microvascular complications (29).

Identifying social determinants affecting people living with diabetes is key to comprehending morbidity contributors and customizing management strategies for diverse populations, recognizing that disadvantaged backgrounds may lead to increased healthcare utilization and emergency care reliance. (30, 31). Issues that affect their health may not be fixed by physical healthcare alone, and a framework of care encompassing a biopsychosocial model may be needed. The Frome model of primary care is a project in Somerset in the UK that leverages existing social networks to improve health outcomes. It works on the basis that health is heavily influenced by social factors and uses community assets such as peer support groups to tackle social determinants of health and provide support to people in the community. This reduced hospital admissions by 14% over a 4 year period (32). We are also at a time period where diabetes technology is ever advancing, and it is imperative that access to technology is equitable across all social classes. Literature from the UK suggests disparities in access to insulin pump and continuous glucose monitoring devices (33). Another study looking at flash glucose monitors showed that “time in range” did not differ between deprivation categories (34). Therefore, it is important to have equitable access to these technologies to allow everyone to benefit.

The objective of this study is to explore the association between area level social deprivation and diabetic kidney disease in a cohort of adults living with diabetes in Ireland. Area based deprivation indices are well established and widely used and facilitate gradients to be demonstrated at a population level (35–39). To our knowledge, this will be the first study in Ireland to look at the association between deprivation and rate of decline in renal function, using a composite, area level measure of deprivation.

## Methods and Analysis

### Methods

This is a retrospective cohort study of people diagnosed with diabetes attending University Hospital Galway, a tertiary referral centre serving a large catchment area in the west of Ireland. We will use an existing dataset from a previous cohort study of people with diabetes, who attended the diabetes centre between 2012 and 2016.

This dataset contains clinical and laboratory data which was obtained from DIAMOND. DIAMOND is the electronic record that is used in University Hospital Galway for people living with diabetes. Demographic data are input by administration staff on registering with the diabetes service. At each clinic visit DIAMOND is then used to record clinical details-anthropometric measures such as blood pressure and weight, medications used, complications and laboratory results.

The remaining laboratory data in the dataset was obtained from the laboratory IT system at University Hospital Galway. This system records longitudinal laboratory measurements on all samples analysed at University Hospital Galway. Longitudinal values for serum creatinine, urine albumin to creatinine ratio (uACR), serum HbA1c, cholesterol, HDL and triglycerides were obtained for each person in the dataset. Isotope dilution mass spectrometry was to measure serum creatinine, conventional Roche Diagnostics assays were used to measure lipids and urine creatinine and high-performance liquid chromatography was used to measure Hba1c.

The dataset contains the datapoints as shown in table 1. It was previously used in a study looking at the prevalence of diabetic kidney disease and rapid renal function decline in adults with diabetes (40).

**Table 1:** Data in existing dataset. ACR= albumin creatinine ratio eGFR= estimated glomerular filtration rate

| Clinical and Demographic Data | Laboratory Data |
| --- | --- |
| Age | Creatinine ($\mu\text{mol/l}$) |
| Gender | eGFR ($\text{mL/min/1.73 m}^2$) |
| Ethnicity | Urine ACR ($\text{mg/mmol}$) |
| Type of Diabetes | HbA1c ($\text{mmol/mol}$) |
| Duration of Diabetes | Cholesterol ($\text{mmol/l}$) |
| Smoking Status | HDL cholesterol ($\text{mmol/l}$) |
| Diabetes Medications Used | LDL cholesterol ($\text{mmol/l}$) |
| Antihypertensive Medications Used | Triglycerides ($\text{mmol/l}$) |
| Body Mass Index (BMI) ($\text{kg/m}^2$) | Urine ACR ($\text{mg/mmol}$) |
| Systolic Blood Pressure ($\text{mmHg}$) | |
| Diastolic Blood Pressure ($\text{mmHg}$) | |

#### Inclusion criteria

-People living with diabetes, attending the diabetes service at Galway University Hospitals with a diagnosis of diabetes (type 1, type 2 and other)

-Over 18 years of age

#### Exclusion criteria

-Insufficient follow up laboratory data (at least 2 values of creatinine >3months apart required for inclusion)

-Insufficient address details to determine address-based deprivation index

-Primary diagnosis of gestational diabetes, impaired glucose tolerance or impaired fasting glucose

The Pobal Haase Pratschke (HP) deprivation index provides a sophisticated indication of deprivation across Ireland (41). It uses small area geography which was developed in 2011 by the Ordnance Survey of Ireland and the Central Statistics office. Small area geography is useful when mapping social and economic data as the areas are homogeneous in their social composition and population size, with a mean of just under 100 households per small area. The HP index maps the overall levels of affluence and deprivation at the level of 18,488 small areas. We will use the index based off the 2016 census as this most accurately reflects deprivation status at the baseline visit for participants in the study. The HP index is constructed based on three dimensions of affluence/disadvantage-demographic profile, social class composition and labour market situation. Demographic profile includes indicators such as level of educational attainment and age of the population, social class composition includes indicators such as type of profession and labour market situation looks indicators such as unemployment rate.

The Pobal HP deprivation index has previously been used in medical research looking at polypharmacy and dependency in older adults and on survival post renal transplant and on chronic dialysis (42–44).

Individual’s addresses will be used to determine their deprivation index using the Pobal HP 2016 deprivation index. This will be done with the assistance of colleagues in the Health Intelligence Unit using Health Atlas Ireland (45). The addresses in the dataset will first be matched to small area ID. This will be done through 2 processes. Firstly, an automated process will be carried out using health atlas Ireland which will match all addresses with a small area ID where possible. Secondly, a manual address matching process will be carried out where the unmatched addresses will be reviewed individually. The addresses will be refined and matched to suggested addresses on health atlas or alternatively searched on the online interactive HP Pobal deprivation map to determine the corresponding small area. The small area will then enable us to determine a deprivation index for each individual in the dataset. These indices are reported as a numerical value from roughly −40(most disadvantaged) to +40(most affluent) or category of relative index score which are defined by HP as per table 2 below. Categories of “extremely” and “very” disadvantaged will be merged into the “disadvantaged” category and similarly for the “affluent” category due to anticipation of low numbers in these categories. For each small area it is also possible to obtain data from which the deprivation scores are constructed such as population change, age dependency ratio, lone parents ratio, education level, unemployment rate, proportion of professional and manual workers, percentage of owner occupied households and rented households and average persons per room.

**Table 2:** Pobal HP deprivation Indices 2016.

| Relative Index Score | Standard Deviation | Label |
| --- | --- | --- |
| Over 30 | >3 | Extremely affluent |
| 20-30 | 2 to 3 | Very affluent |
| 10-20 | 1 to 2 | Affluent |
| 0 to 10 | 0 to 1 | Marginally above average |
| 0 to -10 | 0 to -1 | Marginally below average |
| -10 to -20 | -1 to -2 | Disadvantaged |
| -20 to -30 | -2 to -3 | Very disadvantaged |
| Below -30 | <3 | Extremely Disadvantaged |

For a proportion of the cohort, the address may not be detailed enough to identify a corresponding small area but a larger area may be identified. In these cases, the average deprivation index of each of the small areas within the larger area will be used.

The existing dataset will be updated with serial laboratory measurements of creatinine, urine ACR, Hba1c and lipids until January 2023. eGFR will be calculated using the CKD-EPI 2021 equation (46) for all creatinine values from the last clinical episode date in the existing dataset until January 2023. These eGFR values will then be censored to exclude those on dialysis or who have received renal transplant as these variations in eGFR are not reflective of renal function decline. Rate of renal function decline will be calculated as per the methods in the previous study utilising this dataset (40). Linear mixed-effects models (incorporating random within-subject trajectories of eGFR over time) will be used to generate individual-specific eGFR slopes. These models will be applied to untransformed eGFR measurements to estimate absolute change in eGFR (mL/min/1.73 m^2^/year), and to log-transformed eGFR measurements to estimate percentage change (% change per year). These slopes represent the change in renal function over time for each participant incorporating all eGFR measurements. Only individuals with at least 2 eGFR readings 3 months apart will be included to calculate rate of decline. Progressive or rapid decline in renal function among participants with DM will be defined as either an absolute reduction in eGFR of >3.5ml/min/1.73m^2^/year (47) or proportionate eGFR loss per year of >3.3% (48).

The primary outcome will be rate of renal function decline (absolute and percentage values as per formula above). Secondary outcomes will include time to reaching end stage kidney disease (eGFR<15ml/min/1.73 m^2^), dialysis or renal transplant and variability in eGFR and ACR measurements which may reflect fluctuation in health status which could have been influenced by external social factors.

### Analysis

Stata V.17 will be used for statistical analysis. Data will be assessed for normality. Descriptive analysis will be performed, comparing the distribution of variables between deprivation categories. Age, duration of diabetes, number of antihypertensive medications used, BMI, blood pressure, baseline creatinine, eGFR, uACR, HbA1c and lipids will be described with median/mean with standard deviation values depending on distribution and minimum/maximum values. Frequencies and proportions will be used to describe categorical variables-gender, type of diabetes, ethnicity, smoking status and medications used. ꭓ^2^ squared test will be used to compare categorical variables between deprivation groups. Analysis of variance (ANOVA) will be used to compare means of continuous variables between groups.

Linear regression models will be used to explore the relationship between explanatory variables (including deprivation) and the primary outcome of rate of decline in renal function (absolute and relative based on eGFR). Logistic regression models will be used to explore categories of rapid and non-rapid decline. A “time to event” analysis will be carried out using the endpoints of ESKD/dialysis or renal transplant as described above.

Confounders such as diabetes duration and HbA1cbe adjusted for in the models with careful consideration to differentiate confounders from mediating factors on the causal pathway.

Missing data will be dealt with using a complete case analysis approach. It is anticipated that there will be data missing from the exposure variable (deprivation) as a result of inadequate/incomplete address data and outcome variable (rate of renal function decline) as a result of lack of follow up laboratory data which may be due to change in location of diabetes management or death. Missing data will therefore be excluded from the analysis. A sensitivity analysis will be carried out using only the addresses matched to exact small area (i.e., excluding those with an averaged deprivation index).

A data protection impact assessment (DPIA) has been carried out to ensure the research methodology is in line with general data protection regulation (GDPR). Consent has not been obtained from participants as the data will be anonymised when accessed and analysed and therefore ethical approval was granted without the requirement to obtain consent. All data will be stored on a password protected Health Service Executive laptop with approved encryption software and anti-virus software. The file containing the dataset will be password protected.

The dataset was accessed on 1/12/23 and is currently being updated with the additional laboratory data and deprivation indices as outlined above. The dataset does not contain any information that can identify individual participants. The planned start date for the study analysis is 03/06/2024.

## Results

The baseline characteristics of each individual will be described as per table 3 using the information available from the existing dataset:

**Table 3.**
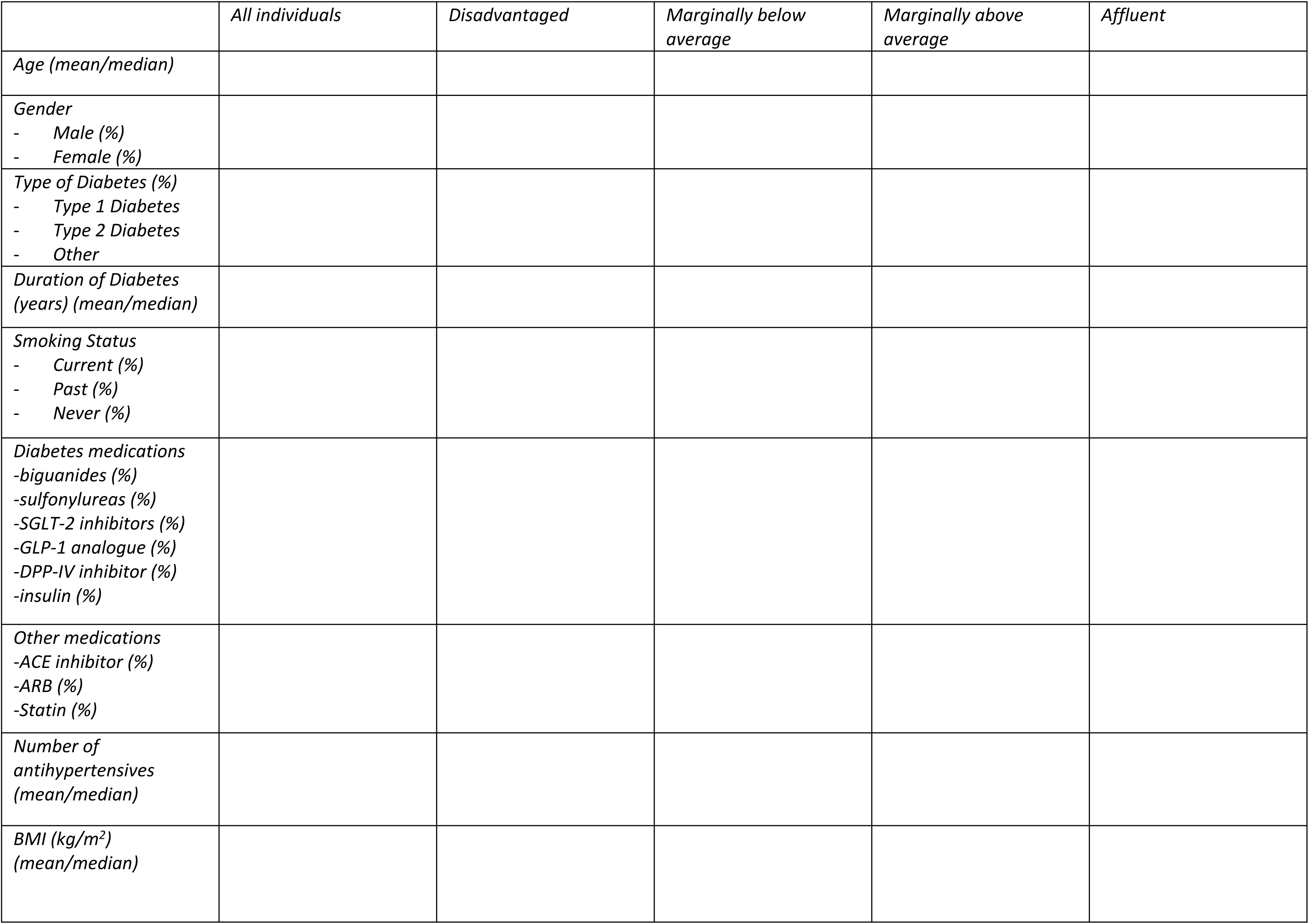
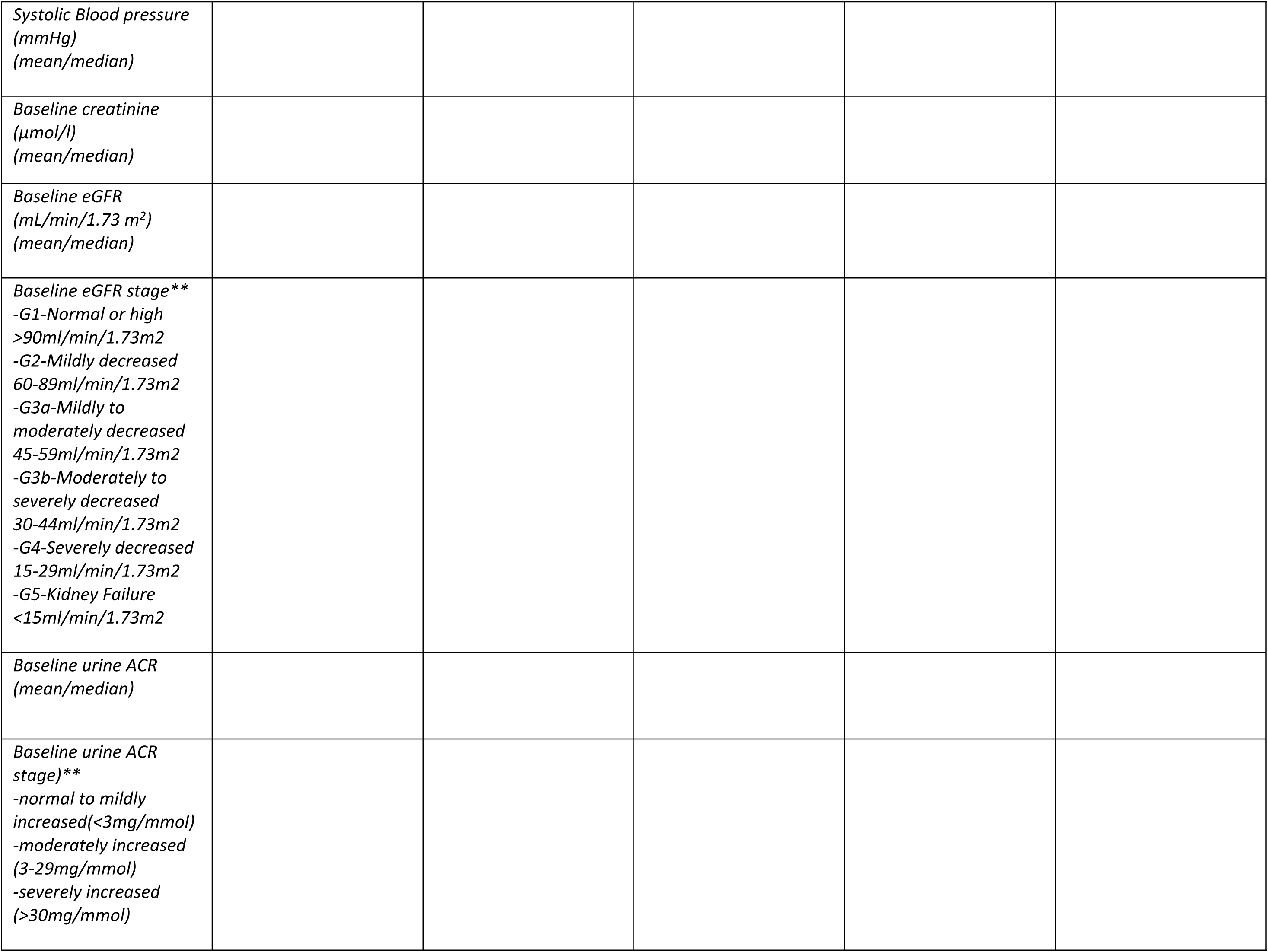
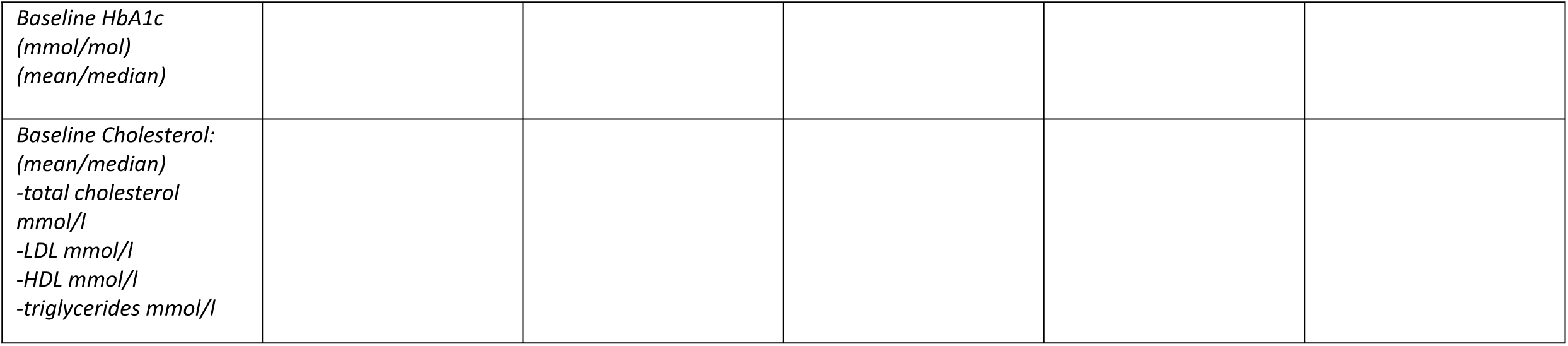
Baseline characteristics of individuals categorised by deprivation status. SGLT-2 inhibitor= sodium-glucose co-transporter-2 inhibitor, GLP-1 agonist= glucagon-like peptide 1 agonist, DPP-IV inhibitor= dipeptidyl-peptidase IV inhibitor, LDL= low-density lipoprotein, HDL= high density lipoprotein, ACE inhibitor= angiotensin-converting-enzyme inhibitors, ARB= angiotensin receptor blocker **Urine ACR and eGFR were classified as per the Kidney Disease: Improving Global Outcomes (KDIGO) 2012 Clinical Practice Guideline

## Discussion

There is a large body of evidence internationally demonstrating that social deprivation has a detrimental outcome on people’s health (49–60). The association between deprivation and diabetes complications has been explored internationally. This study will be the first in Ireland to explore the relationship between area level social deprivation and rate of decline in renal function among people with diabetes. It will be a starting point in a specific geographical area which can then be replicated across the country. If we identify an association between social deprivation and diabetic kidney disease in Ireland, it will highlight a need for targeted interventions for vulnerable subgroups with diabetes. This may be achieved in part through the enhanced community care programme, a national programme in Ireland designed to move the management of chronic diseases (including diabetes) to the community and away from tertiary or hospital care. Delivering care in the community and perhaps targeting more deprived areas could help to address the effect of deprivation. We may need to consider the impact of deprivation at each clinical assessment of a person with diabetes and how we can help modify the effects of same. Our models of care for diabetes management would need to be amended to address the impact of deprivation and how we can address this, for example through equitable access to technology, tailored diabetes education and virtual clinics where appropriate. We would need to ensure that our health system provides equal access to care and ensure there are governmental policies to subsidise healthy foods and provide equitable access to recreational facilities The results of this study may have significant impact on informing new health care and governmental policies.

## Strengths and limitations of this study

- First study in Ireland exploring an association between deprivation and rate of decline in renal function using a composite deprivation index
- Long duration of follow up of longitudinal laboratory data (7+years)
- Follow up results limited to a single tertiary public centre
- Quality of data dependent on accuracy of data entry into electronic patient record system by healthcare professionals

## Ethics and Dissemination

Ethical approval was granted from the Clinical Research Ethics Committee at Galway University Hospitals in March 2023-Ref C.A. 2956. A data protection impact assessment (DPIA) was also completed.

The results will be written up for publication in a peer reviewed journal and presentation at national/and or international diabetes and public health conferences. The findings will be relevant to clinicians managing diabetes, public health specialists and healthcare service managers/policy makers.

## Author Contributions

SD, TG, MG and CC conceived and designed the study. CC wrote the protocol. CMB and PMK refined the study design. All authors revised the protocol.

## Funding Statement

This research received no specific grant from any funding agency in the public, commercial or not-for-profit sectors.

## Competing Interests Statement

CC received funding to attend national and international diabetes conferences from Novo Nordisk. TPG received honoraria for speaker fees and/or advisory boards from Novo Nordisk, Sanofi Aventis, Mundipharma Pharmaceuticals, Dexcom, Abbott Diabetes Care, AbbVie, Astra Zeneca and Eli Lilly.

## Data Availability

Deidentified research data will be made publicly available when the study is completed and published.

## Notes

### Funding Statement

The author(s) received no specific funding for this work.

### Author Declarations

Ethical approval has been granted by the clinical research ethics committee of Galway University Hospitals- Ref C.A. 2956.

